# Understanding the variant landscape, and genetic epidemiology of Multiple Endocrine Neoplasia in India

**DOI:** 10.1101/2023.10.20.23297293

**Authors:** Aastha Vatsyayan, Juhi Bhardwaj, Srashti Jyoti Agrawal, Bhaskar Jyoti Saikia, VR Arvinden, Vigneshwar Senthivel, Suruchi Trehan, Kavita Pandhare, Mohamed Imran, Rahul C Bhoyar, Mohit Kumar Divakar, Anushree Mishra, Bani Jolly, Sridhar Sivasubbu, Vinod Scaria

## Abstract

**Aim:** Multiple Endocrine Neoplasia (MEN) is a familial cancer syndrome that encompasses several different types of endocrine tumors. The disease has three main types, namely MEN1, MEN2 and MEN4 that may or may not overlap phenotypically, but are caused by genetic mutations in three different genes, namely *RET, MEN1* and *CDKN1B* respectively. Genetic testing for effective diagnosis, improved prognosis, and treatment is recommended as part of of clinical practice guidelines, which makes establishment of accurate pathogenicity classification of variants across the three genes essential. However, few resources offer such classification, especially in a population specific manner.

**Materials and Methods:** Using the gold-standard ACMG/AMP guidelines for variant classification, we have systematically classified variants reported across the *RET, MEN1* and *CDKN1B* genes reported in the IndiGen dataset, and established the genetic epidemiology of MEN in the Indian population. We have additionally classified variants from ClinVar and Mastermind, and made all variant classifications freely accessible in the form of a database called MAPVar. Finally, we have designed a primer panel for accurate, cost-effective diagnosis of the three MEN types.

**Results:** We have established the genetic prevalence of MEN in the Indian population to be the following: 1 in nearly 341 individuals is a likely carrier of MEN linked pathogenic *RET* mutations in the Indian population.

We have compiled ACMG-classified variants from three large datasets to create an exhaustive compendium of MEN-linked variants called MEN-Associated Pathogenic Variants (MAPVar). The database is available at: https://clingen.igib.res.in/MAPVar/ We have also designed an NGS primer panel across two pools covering all 33 exonic regions of the three genes through 38 amplicons.

**Conclusion:** Our work establishes that MEN is prevalent disorder in India, with MEN2 variants being the most reported of the three types. This indicates the need of more genomic studies of MEN variants to establish a more comprehensive variant landscape specific to Indian populations.

Additionally, genetic testing is an effective tool used against MEN. Our panel offers a means of swift testing, and the MAPVar resource offers an exhaustive compendium of ACMG-classified MEN variants, that can act as a ready reference to aid in interpretation of genetic testing results, as well as better understanding genetic variants in clinical as well as research settings.

## Introduction

Multiple Endocrine Neoplasia (MEN) is a familial or hereditary cancer syndrome encompassing a group of heterogenous clinical syndromes characterized by the occurrence of two or more endocrine gland tumors in a patient^1^. These syndromes include MEN1, MEN2, and MEN4. All three syndromes are autosomal dominant in nature, and are typically inherited, although sporadic cases are known to occur.

MEN1, caused by inactivating mutations in the *MEN1* tumor suppressor gene, and is typically linked with parathyroid, gastro-entero-pancreatic neuroendocrine, and pituitary tumors. Patients may also develop cutaneous, adrenal cortex, and thyroid tumors, foregut carcinoids and meningiomas. Studies also report that women with MEN1 have a significantly higher risk of developing breast cancer.

MEN2 is caused by activation mutations in the *RET* proto-oncogene, and is further divided into three subtypes based on the disease phenotype: MEN2A, MEN2B and familial medullary thyroid carcinoma (FMTC). MEN2A typically presents with medullary thyroid carcinoma (MTC) and pheochromocytoma, while MEN2B additionally presents with manifestations such as “Marfanoid” body habitus, multiple mucosal neuromas, protruding lips, bone abnormalities, corneal nerve thickening etc. MEN2B patients exhibit more aggressive and penetrative MTC, but generally do not develop parathyroid disease. FMTC usually involves only MTC, and is considered a mild version of MEN2A.

MEN4 presents similarly to MEN1, but is caused by inactivating mutations in *CDKN1B* gene. The age of onset of MEN4 is later than MEN1, and the incidence of gastro-entero-pancreatic endocrine tumors is lower. Pituitary tumors are also less common in MEN4. Figure 1 summarizes the different types of MEN disease.

**Figure 1:**
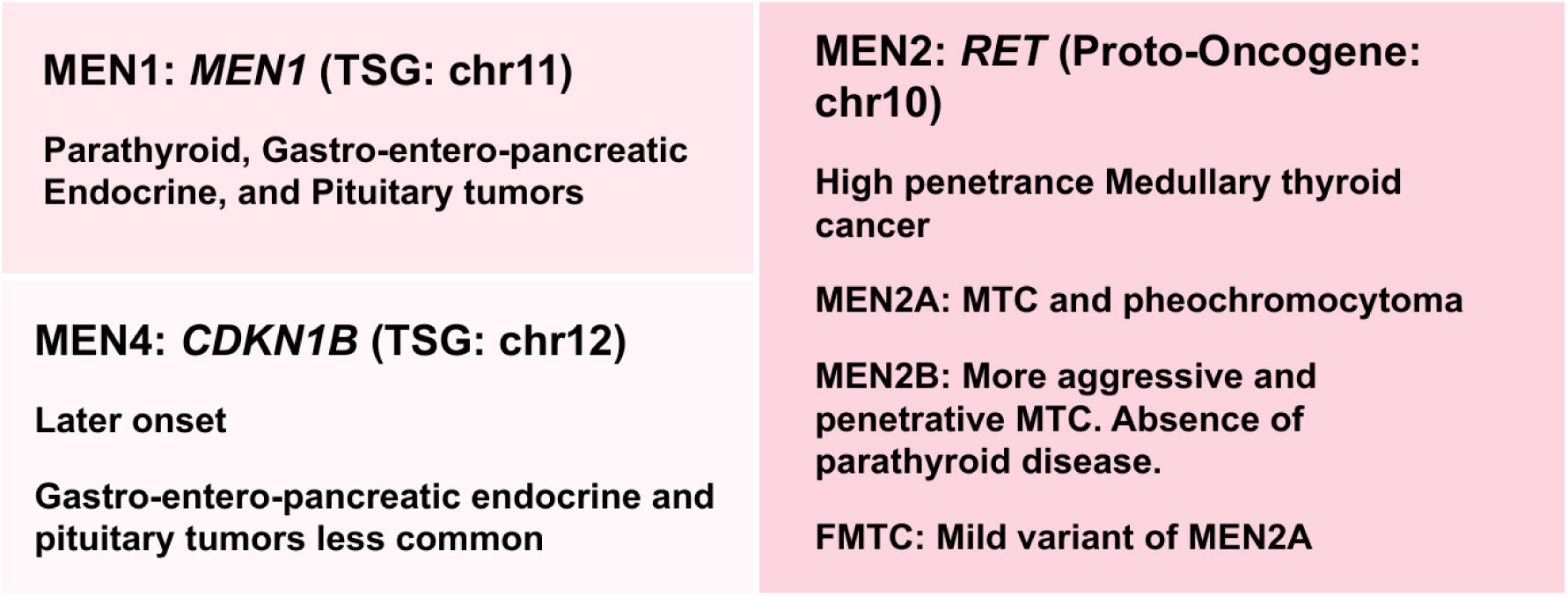
Summary of the different types of MEN disorders classified on the basis on their causative gene

While MEN is linked with high penetrance (93-95%), and presents at a young age (30 - 50 years)^2^ it remains an under-studied familial cancer syndrome in India, with outdated prevalence numbers reported only in global populations. With creation of IndiGen^3^, the population-scale whole-genome sequencing dataset from 1029 healthy individuals from across the country, it is now possible to estimate and establish the genetic prevalence and variant landscape of variants linked with MEN in Indian populations. Essential to this endeavor are the variant classification guidelines provided by the American College of Medical Genetics and Genomics (ACMG) along with the Association for Molecular Pathology (AMP).

In this study, we systematically classify variants linked with MEN across the *RET*, *MEN1*, and *CDKN1B* genes reported in the IndiGen dataset in order to establish the genetic epidemiology of MEN in the Indian populations. We additionally classify variants in the three genes reported across 2 other large-scale datasets, namely ClinVar^4^ and Mastermind^5^, and create a compendium called MEN-Associated Pathogenic Variants (MAPVar) for ready reference to be utilized in clinical as well as research settings. We also prove the utility of the database by querying the GUaRDIAN^6^ cohort data, a nation-wide collaborative framework for decoding rare diseases in India. With a total of 3,280 ACMG-classified variants, the MAPVar database is accessible at: https://clingen.igib.res.in/MAPVar/

## Materials and Methods

### Data collection and variant annotation

We queried three large datasets for all reported variants across the *RET*, *MEN1* and *CDKN1B* genes; these included all variants reported in Mastermind and IndiGen, as well as all ClinVar VUS variants. We annotated all collected variants using the ANNOVAR^7^ tool (ver 2018-04-06), which utilized several databases to annotate variants with positional information (RefGene^8^ database), details regarding protein change (dbSNP^9^), as well as allele frequencies from global population datasets including gnomAD^10^ v3, the 1000 Genomes Project^11^ (1KG), Esp6500^12^ and Greater Middle East^13^ (GME). It also added computational predictions and pathogenicity scores generated by benchmarked tools including SIFT^14^, PolyPhen2^15^, and CADD^16^.

We further queried the ClinVar database programmatically, and annotated the variants with the clinical significance reported there. We removed all variants reported as ClinVar benign/likely benign, as well as all non exonic variants, and also filtered in all Mastermind variants reported as being high-confidence variants reported in literature.

### ACMG/AMP classification and establishment of genetic epidemiology

The variants for the three genes thus obtained were classified using the ACMG/AMP guidelines. Each variant was individually assessed for all 28 attributes that included variant allele frequencies reported across different populations, pathogenicity prediction scores from several tools, variant annotations obtained from various databases, and a thorough literature survey of studies performed on each variant. A detailed explanation of each attribute is provided in Supplementary Data 1. The attributes that were thus assigned to a variant were tallied using the Genetic Variant Interpretation Tool^17^ which generated the final classification for the variant.

Using the pathogenic mutations obtained in the IndiGen dataset, we established the genetic epidemiology of MEN in India.

### Comparison with other populations

Upon classification, we collected all pathogenic/likely pathogenic variants obtained and compared their allele frequencies reported across 18 global populations datasets. These included 1KG, gnomAD, China Metabolic Analytics Project^18^ (ChinaMAP), the Hong Kong Cantonese population^19^ (HKG), TogoVar^20^ (Japanese population), Korea1K Variome^21^ (KGP), Korean Variant Archive 2^22^ (KOVA 2), Qatar^23^, Taiwan Biobank^24^, GME^13^ populations and subpopulations, the Gambian^25^ dataset (Gambian), the GenomeAsia100K Project^26^ (GenomeAsia), Human Genome Diversity Project^27^ (HGDP), the Andamanese population^28^ (Andamanese), the Simons Genome Diversity Project^29^ (Simons), the Singapore Sequencing Indian Project^30^ (SSIP), the Singapore Sequencing Malay Project^31^ (SSMP), and the Iranome^32^.

### MAPVar database and web interface design

We compiled all variants classified from across the three databases, along with the annotations used for ACMG classification, as well as the final classification category of the variant into the MAPVar database.

The database interface was equipped with a search panel that enables users to access data through a wide variety of search queries including variant ID, nucleotide change, amino acid change, as well as rsIDs from dbSNP. The data was ported onto MongoDB (ver. 3.4.10), an open-source NoSQL database system, after being transformed into JavaScript Object Notation format. The web interface is running on Apache HTTP server, using PHP 7.0. The web interface was coded in PHP, AngularJS, HTML, CSS and Bootstrap4.

### Patient Validation

We queried the pathogenic / likely pathogenic variants obtained in MAPVar database across the GUaRDIAN data, which encompasses multiple rare disease cohorts from across the country.

### Multiplex primer designing

We designed a panel for *RET, MEN1* and *CDKN1B* genes to enable quick detection. Multiple primers were designed to cover all coding regions of the three genes. The Primer3plus^33^ along with Oligocalc^34^ were used to select ideal primer length, melting temperature (Tm), as well as GC content. The ThermoFisher Multiple Primer Analyzer^35^, and PrimerPooler^36^ tools were further used ensure that the panel pools were accurately chosen and functioned as expected.

### Sequencing and Bioinformatic Analysis

A cohort of 72 patients afflicted from a myriad of endocrine disorders was sequenced in order to assist in diagnosis. DNA was extracted from patient samples using the salting out method^37^ developed by Miller et. al. The desired regions were amplified using PrimeSTAR GXL DNA Polymerase (TAKARA Bio. Inc, Japan). The mastermix prepared is described in Table 1.

**Table 1:**
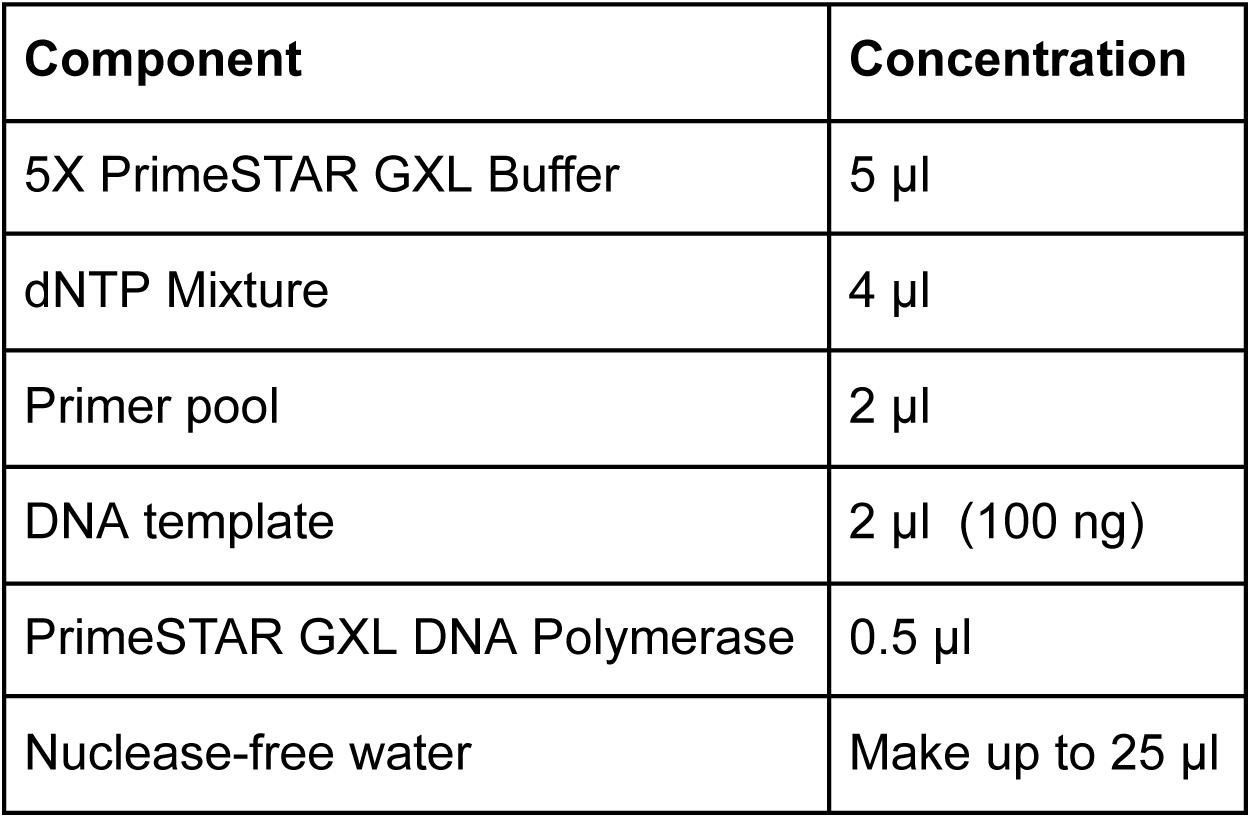
The preparation of the PCR mastermix.

| Component | Concentration |
| --- | --- |
| 5X PrimeSTAR GXL Buffer | 5 µl |
| dNTP Mixture | 4 µl |
| Primer pool | 2 µl |
| DNA template | 2 µl (100 ng) |
| PrimeSTAR GXL DNA Polymerase | 0.5 µl |
| Nuclease-free water | Make up to 25 µl |

The amplification was then performed using the conditions shown in Table 2 in the thermal cycler. The amplified PCR products were visualized by agarose gel electrophoresis 1.2% (w/v) and using the Bio-Rad gel documentation system.

**Table 2:**
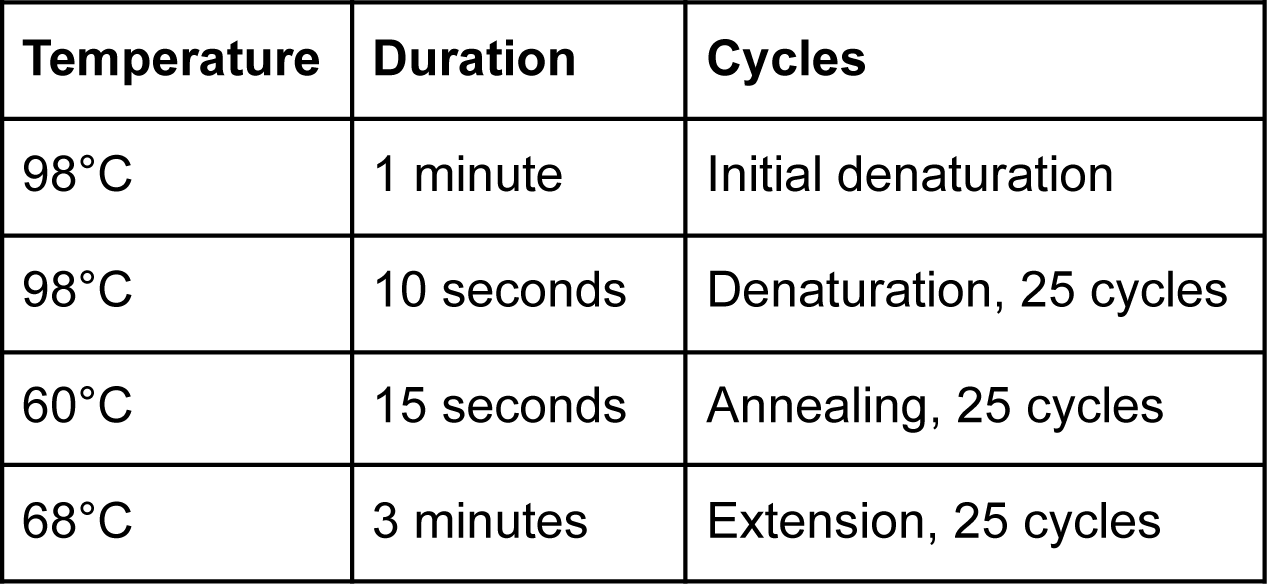
Conditions set in the Thermal cycler for amplification.

The amplified PCR products were then tagmented (tagged and fragmented), and then indexed and amplified to become sequencing-ready libraries. The final libraries were visualized on agarose gel (1.8% w/v). Paired-end sequencing was then performed using the Illumina NGS platform NovaSeq.

The resulting BCL files were then demultiplexed using Illumina’s bcl2fastq^38^ tool. FastQC^39^ and Trimmomatic (v0.39)^40^ tools were then used to perform quality control on the resulting fastq files, which were then aligned to the hg38 human reference genome using the BWA-MEM (v0.7.17)^41^ tool. The aligned files are next converted to BAM format and sorted using SAMtools (v1.13)^42^. Picard’s^43^ MarkDuplicates utility (v2.4.1) was then used to remove duplicate reads from the BAM files. Finally, the BAMs were converted into pileup using the SAMtools mpileup utility, which was then input through VarScan^44^ (v2.4.4) for variant calling. Downstream tools such as ANNOVAR and Ensembl Variant Effect Predictor (VEP)^45^ were then used to annotate the variants with RefSeq^46^ information describing the variant in greater detail, as well as clinical actionability of each variant.

## Results & Discussion

### ACMG/AMP classification and establishment of genetic epidemiology

A total of 4,477 *RET, MEN1* and *CDKN1B* variants were obtained from ClinVar, 1,369 from Mastermind, and 1,787 from IndiGen databases respectively. The collected variants were processed using bash scripts to obtain their coordinates as per the GRCh38 genome assembly; the ANNOVAR tool was then run to annotate the variants with gene name, allele frequencies and pathogenicity scores. Scripts were then used to annotate the variants with clinical significance from ClinVar. The resulting exonic variants across the three genes that were not reported as benign/likely benign in ClinVar were filtering in. A total of 1,324 unique variants were thus obtained. All the variants were systematically classified according to the ACMG/AMP guidelines.

Upon final classification using the Genetic Variant Interpretation Tool, all pathogenic variants across the three genes reported in the IndiGen dataset (Table 3) were used to calculate the genetic prevalence of MEN in India, using the following formula:

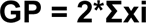

**Table 3:** Pathogenic *RET* variants in the IndiGen data obtained after application of quality cut-offs and classification through ACMG guidelines.

| ID | Variant | rsID | Gene | Attributes | ACMG Classification |
| --- | --- | --- | --- | --- | --- |
| 10:43114507:C:T | RET:NM_020975:exon11:c.1907C>T:p.T636M | rs1035958105 | RET | PM2, BP4, PS3 | Likely pathogenic (II) |
| 10:43102608:G:A | NM_020975:exon3:c.604G>A:p.V202M | rs751572082 | RET | PM5, PM2, PP3, PM1 | Likely pathogenic (IV) |

where, GP = Genetic Prevalence, xi = allele frequency of pathogenic variants

We determined that 1 in nearly 341 individuals is a likely carrier of MEN linked pathogenic *RET* mutations in the Indian population.

It is important to note here that the second variant has not been strongly classified as being likely pathogenic (i.e. it is not Likely pathogenic (I) or (II)), as it relies solely on non-functional supporting data for variant classification. A highly stringent approach could involve the removal of such variants from prevalence estimate calculations. We have included the variant however, as no stringency cutoffs currently exist for such calculations.

The first variant is present in an individual from Jammu and Kashmir, while the second variant is present in two individuals from Odisha. These variants, especially in people from the two states, should be included in screening efforts. Additionally, they offer two important targets for further prevalence studies in Indians, and functional studies for establishment and understanding of the mechanism of action involved.

### Comparison with other populations

We compared all the likely pathogenic variant we obtained from the IndiGen database in the RET gene with 18 population-scale datasets. We did not find the first variant across any population. The second variant was found at low allele frequencies (AF) in ChinaMAP [AF=0.00004722], gnomAD global [AF=0.0000591], gnomAD AMR[AF=0.000588], gnomAD AFR [AF=0.0000241], and gnomAD NFE [AF=0.000102] populations.

Upon performing Fisher’s Exact Test of significance, we discovered that at p-value < 0.05, the variant was present at significantly different allele frequencies across all population datasets with respect to IndiGen, where it is present at a comparatively higher allele frequency of 0.000977.

### The MAPVar database

We obtained a total of 1,324 unique variants across *RET*, *MEN1* and *CDKN1B* genes from across IndiGen, ClinVar and Mastermind databases. We processed and annotated this data, and compiled it into the MAPVar database to act as a powerful reference resource for understanding the pathogenicity of MEN-linked variants. We additionally added all variants that had been annotated as per ACMG guidelines by a ClinVar expert panel to the database.

Figure 2 captures the database interface.

Thus, in all, we obtained 1094 Pathogenic/Likely pathogenic, 1591 Benign/Likely benign, and 595 VUS variants, bringing the database to a total of 3280 ACMG-classified MEN-linked variants. Figure 3 shows the gene-wise breakdown of the database numbers.

**Figure 2:**
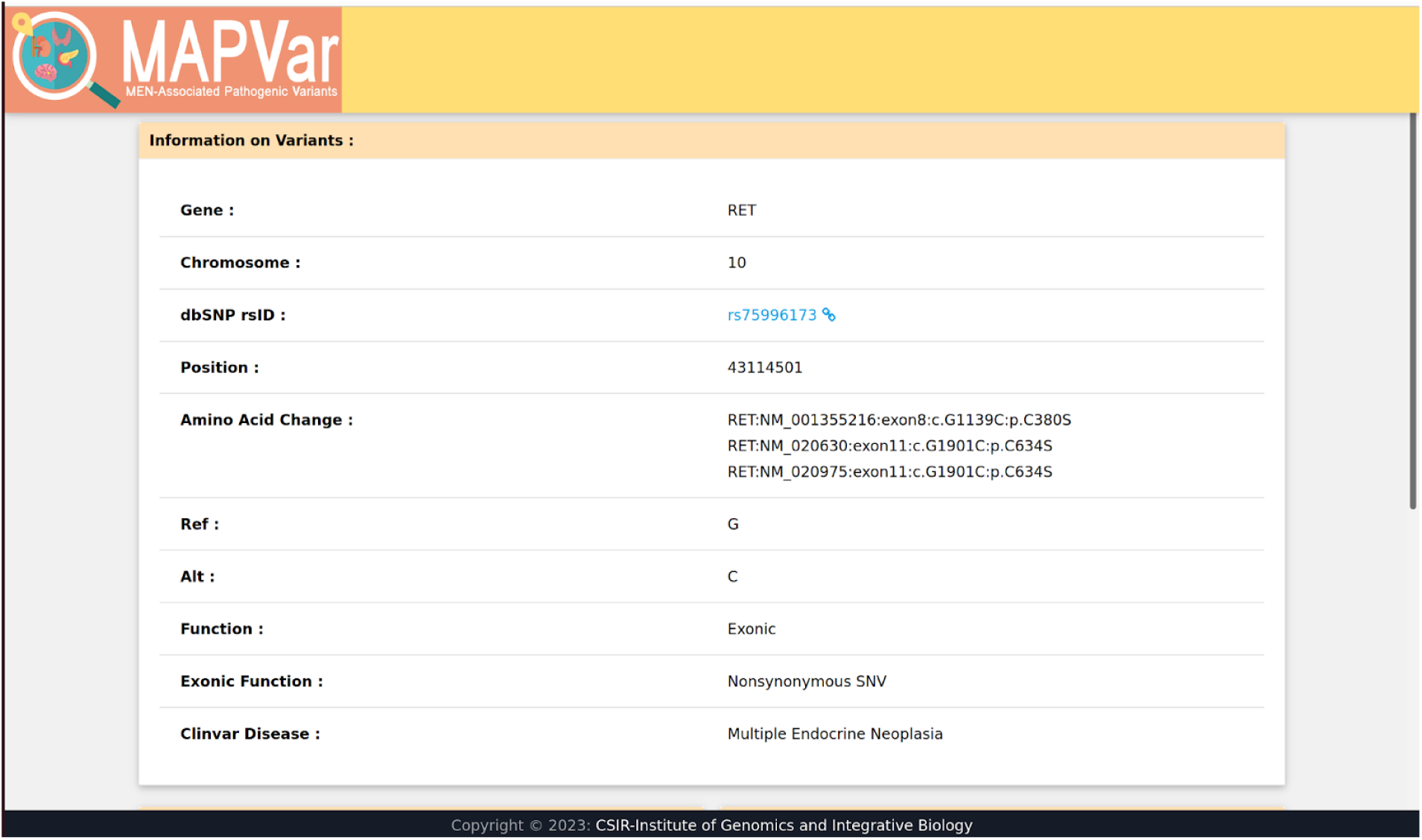
The interface of the MAPVar database, a compendium of MEN linked pathogenic variants

**Figure 3:**
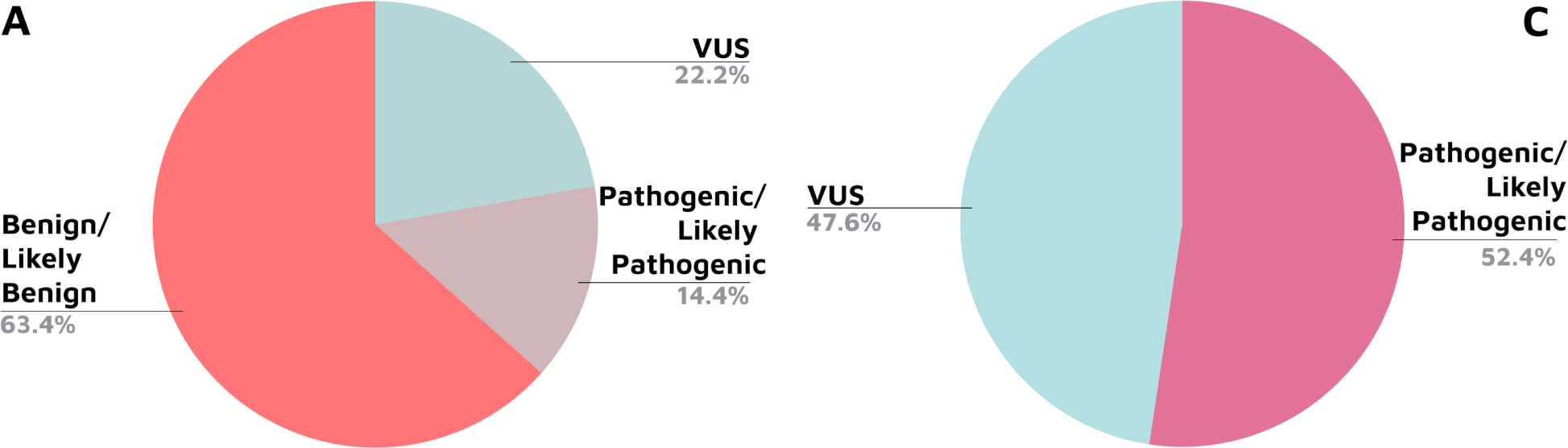
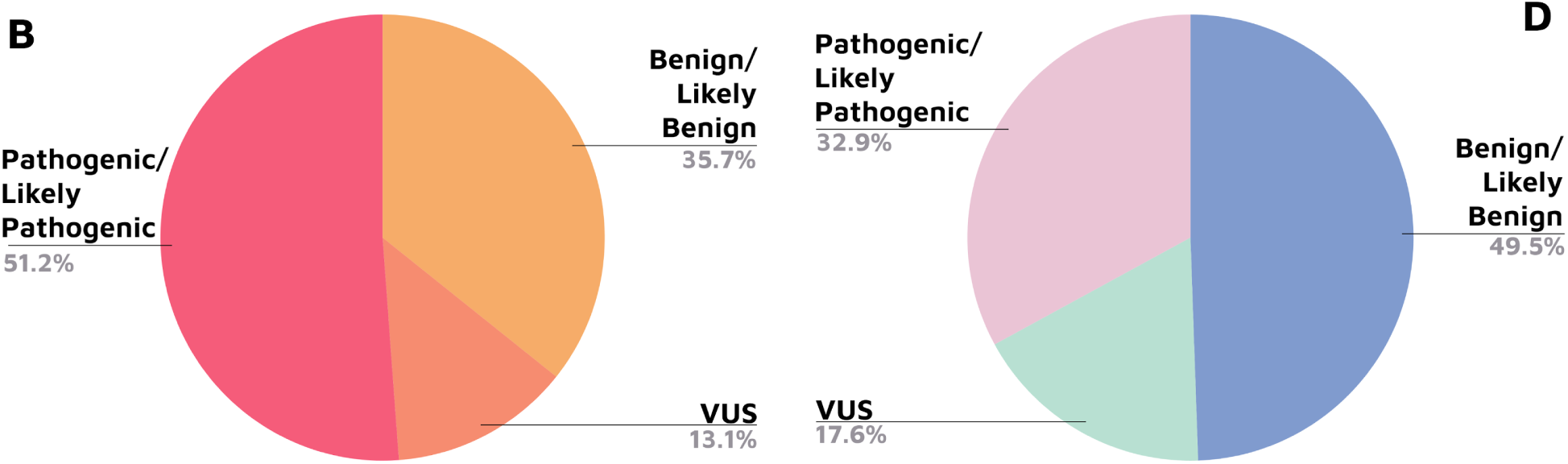
Gene-wise breakdown of the MAPVar database numbers. Panel A: *RET* gene, Panel B: *MEN1* gene, Panel C: *CDKN1B* gene, Panel D: Overall MAPVar numbers.

The database offers multiple formats to query the database with, all of which are enlisted on the homepage. Once a query in any of the enlisted formats is entered, a box containing the preliminary search results is displayed. Upon clicking on the “More Information” button in the box on the desired result entry, a new page bearing detailed information about the variant is opened. This includes information describing the variant, such as gene, chromosome, amino acid changes across various transcripts, alleles, as well as function of the gene and the disease it’s linked with. It also displays the allele frequencies across three global population sets (gnomAD, 1KG, and Esp6500), along with computational predictions made by SIFT^14^, PolyPhen2^15^ and CADD^16^. Finally, the ACMG attributes associated with each variants, and the final classification obtained for the variant are provided in a separate section. Figure 4 shows the detailed search results page of the database, obtained upon clicking the “More Information” button in the expanded search results section of the homepage.

**Figure 4:**
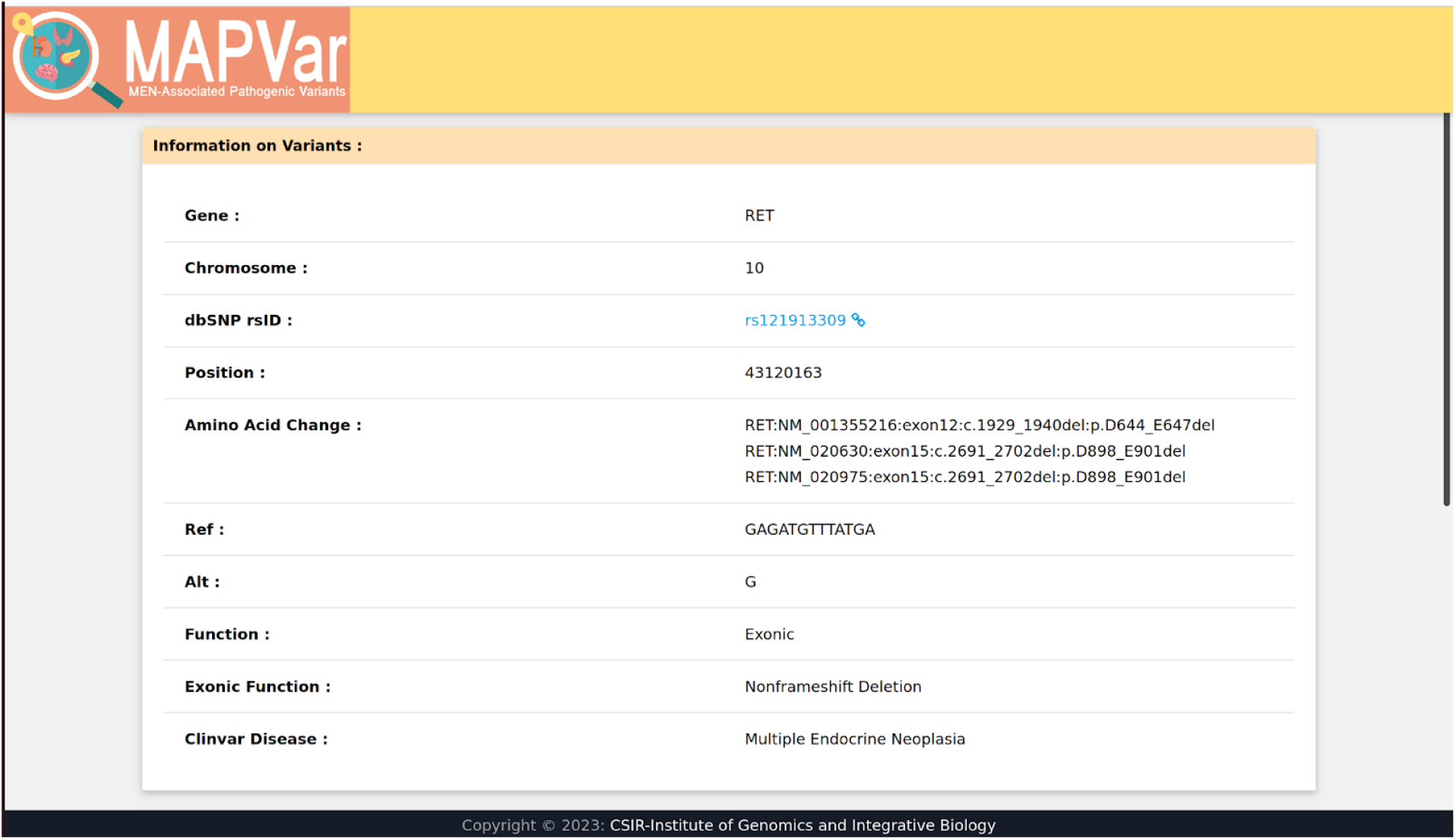
The detailed search results page of the database

MAPVar is accessible at: https://clingen.igib.res.in/MAPVar/

### Patient Validation

Using the MAPVar dataset, we obtained hits for 7 seven patient cases through the endocrine cohort of the GUaRDIAN project data. The cases encompassed 4 provisional diagnoses of MEN, and 3 suggestive of pheochromocytoma, which is often observed in MEN Type 2. A total of 5 unique variants were observed.

Table 4 shows the details of the variants.

**Table 4:**
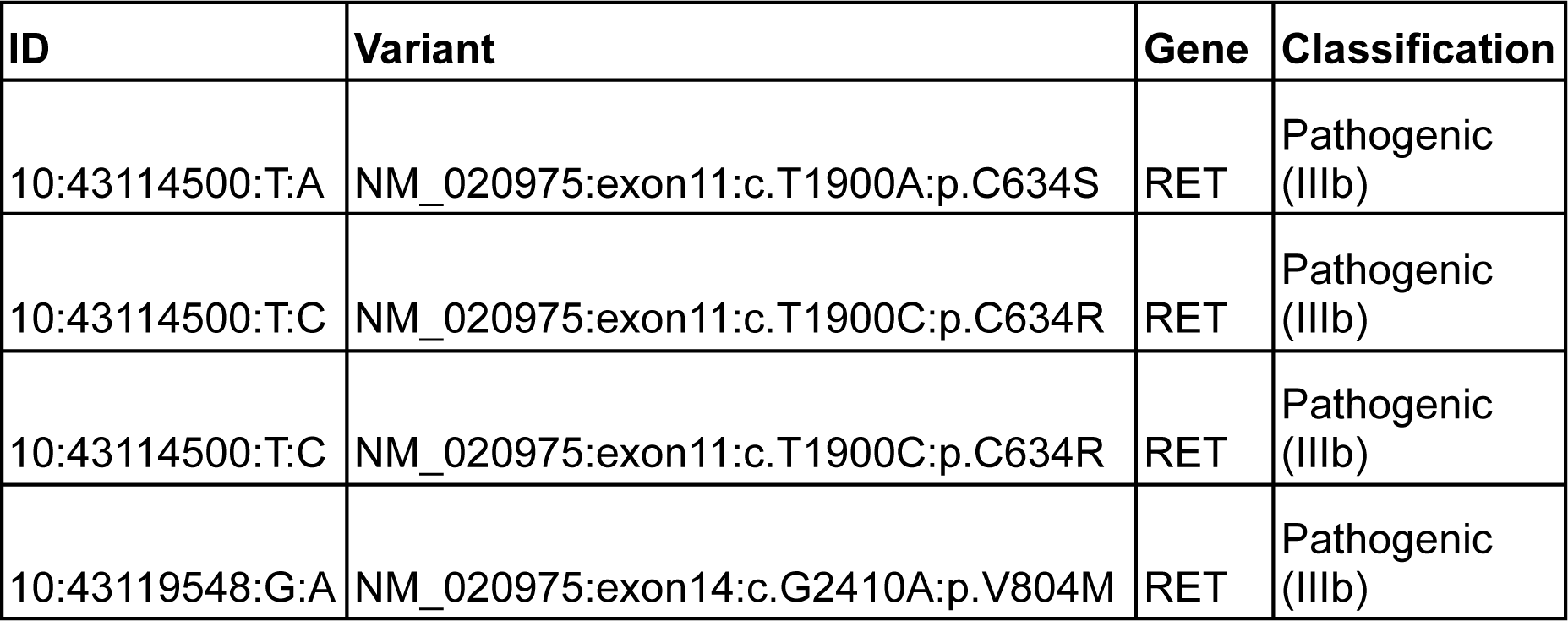
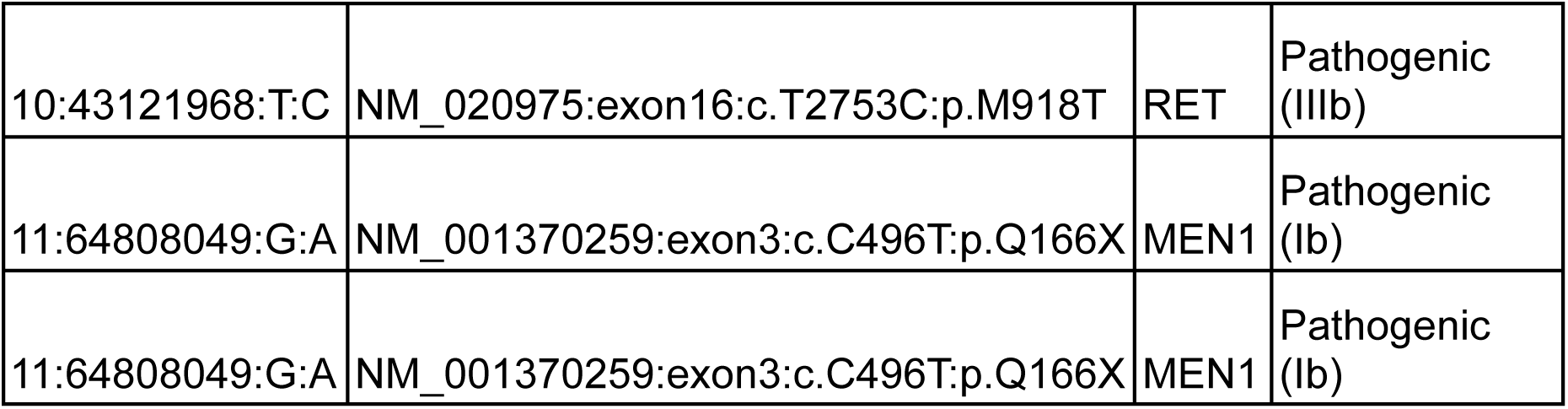
The pathogenic variants found that were reported across the GUaRDIAN database’s endocrine cohort.

### Multiplex primer panel

We successfully created a panel covering all coding regions of *RET, MEN1,* and *CDKN1B* genes. The panel covers 38 amplicons divided into 2 pools, with a total amplified region of 65kb across 33 exons. Figure 5 shows the log2 of average mean coverage across all 72 samples of all three genes.

**Figure 5:**
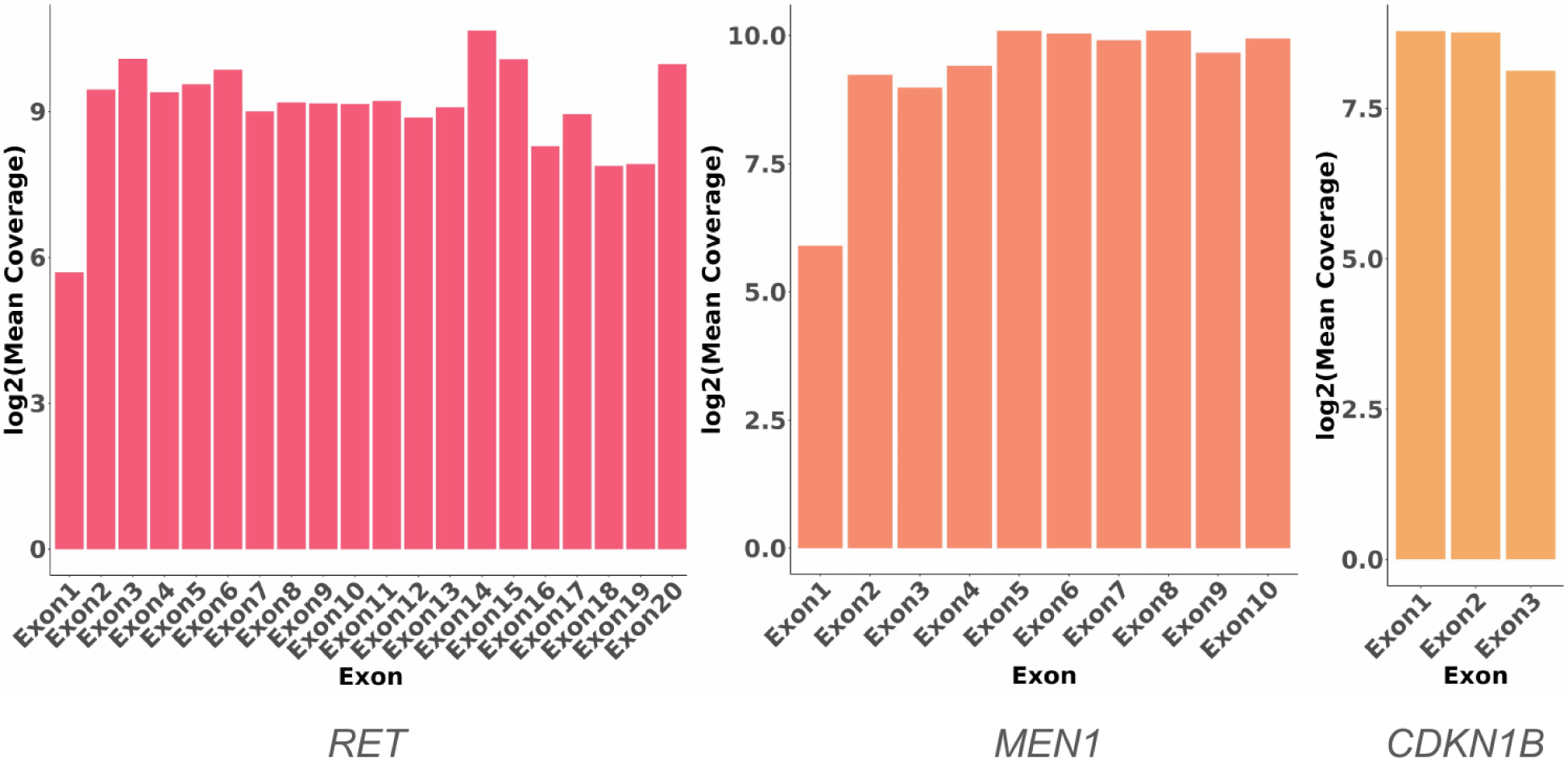
The log2 of exon-wise mean coverage across the three MEN genes in the panel

The primer sequences designed and standardized are detailed in Table 5. Additional positional (hg38), temperature and GC details are shown in Table 6.

**Table 5:**
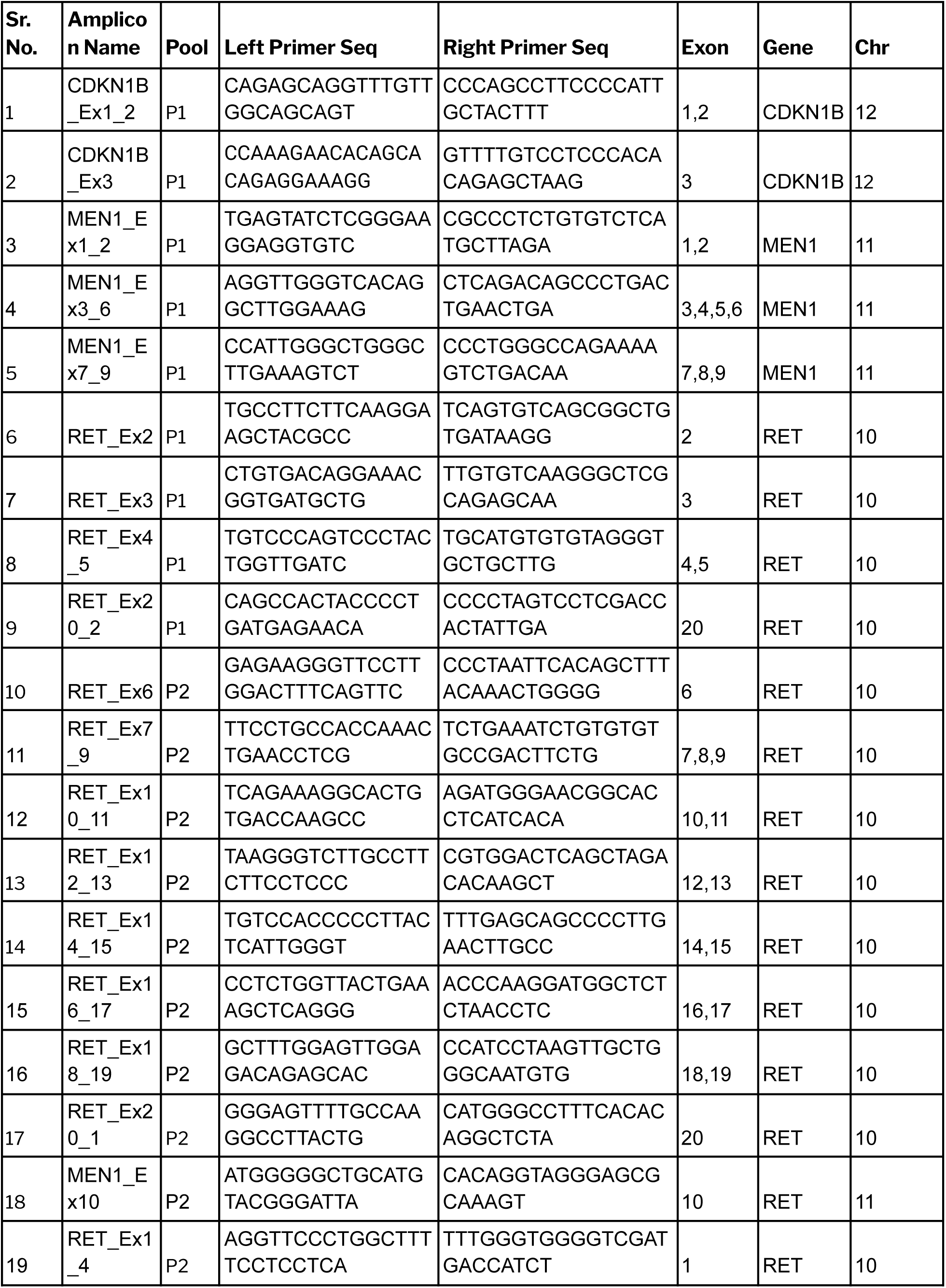
A list of the primers used in the panel for the swift and accurate determination of carrier status, as well as diagnosis of MEN.

**Table 6:**
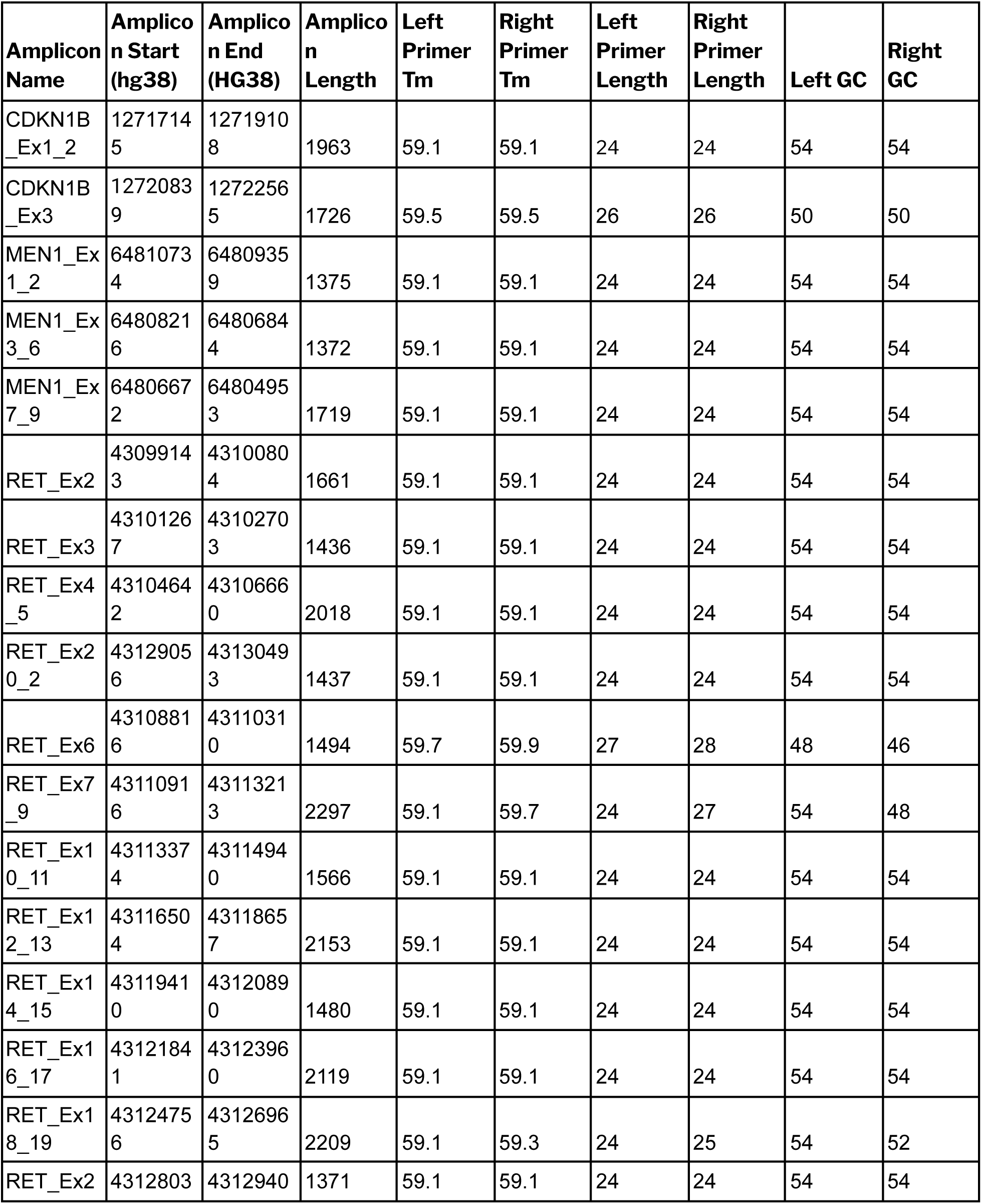
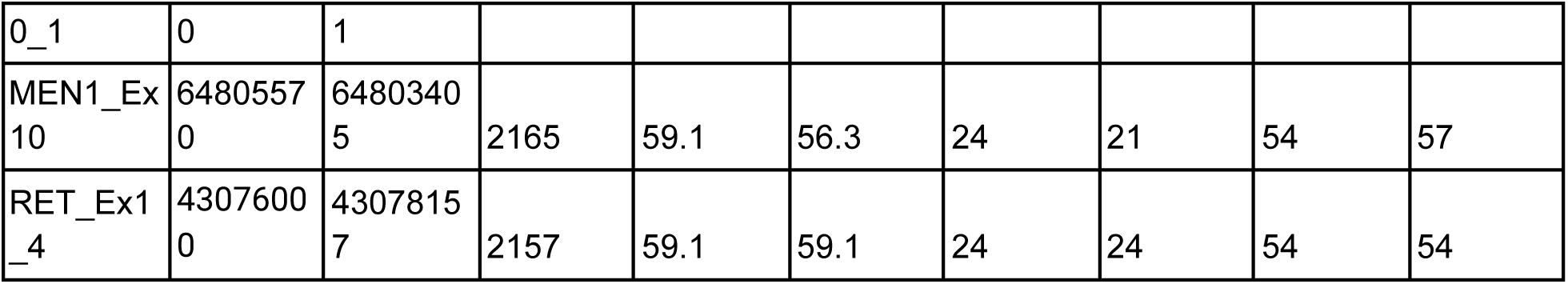
Details of the primers designed for the MEN panel, including position (hg38), primer length, melting temperature (Tm), as well as GC content.

Using this panel, we have successfully been able to solve 3 cases in a cohort of mixed endocrine cancer related cases, where the mutations were known to be pathogenic in nature. The others are being classified through the use of ACMG guidelines. Table 7 depicts the details of the 3 cases.

**Table 7:** Details of cases in which the causative pathogenic mutation has been successfully.

| Patient Variant | Gene | Provisional Diagnosis | Diagnosis Indicated |
| --- | --- | --- | --- |
| MEN1:NM_001370259:exon4:c.C778T:p.Q260X | *MEN1* | Acromegaly | MEN1 |
| MEN1:NM_001370259:exon6:c.912+1G>A | *MEN1* | Pheochromocytoma | MEN1 |
| MEN1:NM_001370259:exon2:c.301dupG:p.V101Gfs*16 | *MEN1* | Insulinoma | MEN1 |

## Discussion

Genetic testing forms an important part of MEN diagnosis. Clinical practice guidelines recommend genetic diagnosis of the three MEN^1^ genes to identify the type of MEN disease. Further, it can be used to differentiate between MEN1 and MEN4, both of which present with similar clinical manifestations, but may result in a different disease prognoses^47^. Guidelines also state that presymptomatic tumor detection and undertaking treatment specific for MEN1 tumors could lead to an improved prognosis in MEN1 patients^48^. Patients diagnosed with MEN are further recommended to have their blood-related family members tested.

Apart from early diagnosis and identification of asymptomatic carriers, genetic testing has also been shown to aid in preimplantation diagnosis^1,49–51^, thereby offering yet another way of tackling MEN cancers preemptively and more effectively. Thus, genetic testing forms an integral part of several stages of the MEN clinical process, and through our work we have aimed to facilitate it.

## Conclusions

Multiple Endocrine Neoplasia remains an understudied cancer syndrome in the Indian population. However, in our work we have established that MEN, especially linked with the *RET* gene is highly prevalent, and each gene, especially *CDKN1B* deserves more studies to establish the pathogenic variant landscape specific to India.

Further, through our NGS panel, we aim to enable swift and cost effective genetic testing of MEN, which would aid and assist in early and accurate diagnosis. Our database then offers a powerful ready resource to access classification of variants linked with MEN, to enable easier interpretation of genetic testing reports, and also to understand variants in clinical as well as research settings better. The results obtained through the analysis of the GUaRDIAN endocrine cohort further prove the utility of the dataset in accurately identifying MEN-linked variants in patients.

Thus through the use of the genetic test as well as the resource for easy interpretation, we hope to be able to address the MEN linked cancer burden in a meaningful manner.

## Acknowledgements

Authors acknowledge funding from the Council of Scientific and Industrial Research (CSIR) through CNP-0007 Grant. The funders had no role in the preparation of the manuscript or decision to publish.

## Author Contributions

VS conceptualized, designed and supervised the study.

AV performed ACMG classification, Epidemiology calculation, all other analyses, and complied the manuscript.

KP created the database interface.

RCB, MI, VS, MKD, AM and BJ were involved in the generation of the study data.

MI and AVR performed the primer standardization and sequencing, and bioinformatics analysis respectively.

VS and SSB conceived and designed the project.

All other authors assisted with the ACMG classification. All authors have read and approved the final manuscript.

## Data Availability

All data produced are available online at: https://clingen.igib.res.in/MAPVar/

